# Comparing safety, performance and user perceptions of a patient-specific indication-based prescribing tool with current practice: A mixed-methods randomised user testing study

**DOI:** 10.1101/2024.07.01.24309757

**Authors:** Calandra Feather, Nicholas Appelbaum, Jonathan Clarke, Ara Darzi, Bryony Franklin

## Abstract

**Background:** Medication errors are the leading cause of preventable harm in healthcare. Despite proliferation of medication-related clinical decision support systems (CDSS), current systems have limitations. We therefore developed an indication-based prescribing tool. This performs dose calculations using an underlying formulary and provides patient-specific dosing recommendations. Objectives were to compare the incidence and types of erroneous medication orders, time to prescribe (TTP), and perceived workload using the NASA task load index (TLX), in simulated prescribing tasks with and without this intervention. We also sought to identify workflow steps most vulnerable to error and gain participant feedback.

**Methods:** A simulated, randomised, cross-over exploratory study was conducted at a London NHS Trust. Participants completed five simulated prescribing tasks with, and five without, the intervention. Data collection methods comprised direct observation of prescribing tasks, self-reported task load and semi-structured interviews. A concurrent triangulation design combined quantitative and qualitative data.

**Results:** 24 participants completed a total of 240 medication orders. The intervention was associated with fewer prescribing errors (6.6% of 120 medications) compared to standard practice (28.3%; relative risk reduction 76.5% p < 0.01), a shorter TTP and lower overall NASA TLX scores (p < 0.01). Control arm workflow vulnerabilities included failures in identifying correct doses, applying maximum dose limits, and calculating patient-specific dosages. Intervention arm errors primarily stemmed from misidentifying patient-specific information from the medication scenario. Thematic analysis of participant interviews identified six themes: Navigating trust and familiarity, addressing challenges and suggestions for improvement, integration of local guidelines and existing CDSS, intervention endorsement, ‘search by indication’ and targeting specific patient and staff groups.

**Conclusion:** The intervention represents a promising advancement in medication safety, with implications for enhancing patient safety and efficiency. Further real-world evaluation and development of the system to meet the needs of more diverse patient groups, users and healthcare settings is now required.

*What is already known on this topic?:* Indication-based prescribing has the potential to improve prescribing efficiency and patient safety.

*What this study adds:* An indication-based, patient-specific prescribing tool used in a simulation setting reduced the incidence of prescribing errors and the time to prescribe compared with standard practice. This study provides cumulative validity to the potential benefits of indication-based prescribing tools.

*How this study might affect research, practice or policy:* Future evaluation of such tools in the real-world clinical setting is now required to identify the impact of such tools on clinical outcomes and prescribing workflow.

## Introduction

Medication errors are the leading cause of preventable harm in healthcare settings worldwide (1). An estimated 237 million medication errors occur in England alone every year, with 66 million considered clinically significant (2). Avoidable adverse drug events related to these errors are estimated to cost the NHS in excess of £98.5 million per year, consuming 181,626 bed-days and causing 712 deaths (2).

Medication-related clinical decision support systems (CDSS), often integrated with electronic prescribing (eP), have proliferated over the last few decades. Functionality provided by these systems has typically been limited to alerts pertaining to drug-drug interactions, allergies, duplications and basic dose range checking. A recent systematic review found such systems to be relatively immature, with little to no human factors input during development and functionality largely generic to all patients (3). There is therefore an opportunity to improve CDSS for patient safety using human factors and usability engineering to ensure systems are both user-friendly and safe for their intended use (4).

A US study of an indication-based prescribing tool demonstrated improvements to efficiency, medication error rates and user satisfaction (7). However, evaluation of similar interventions in different contexts, with different systems and users is required. Our objectives were to compare the incidence and types of erroneous medication orders, time to prescribe, and perceived workload, in simulated prescribing tasks with and without the use of a patient-specific, indication-based prescribing intervention. In addition, we sought to identify workflow steps most vulnerable to error and to gain participant feedback regarding use of the intervention.

## Methods

### Study design and setting

We used a simulated, randomised, cross-over exploratory study to compare prescribing with and without use of a prototype CDSS at a large London teaching NHS Trust, from December 2022 to April 2023. Both quantitative and qualitative methods and analysis were utilised in a concurrent triangulation design, whereby a combination of methods and outcomes can provide an expanded understanding of the studied phenomena (8,9) (supplementary Figure 1). The trust has a range of medical and surgical specialities treating patients of all ages and utilises Cerner (10) as its primary electronic heath record and eP system.

### The intervention

Touchdose is an on-demand clinical decision support tool that receives medication, indication and patient inputs and uses them to apply clinical logic to an underlying formulary, primarily the British National Formulary (BNF) (11) and the BNF for Children (BNFc) (12). Touchdose, a UK conformity-assessed medical device, integrates with the electronic health system and performs dosing calculations as needed, to return patient-specific, indication-based dosing recommendations.

### Identification of participants

Participants were a convenience sample of clinicians who were approached if they regularly prescribed medications for hospital inpatients. Targeted sampling was used to recruit both medical and non-medical prescribers, from a wide range of specialities and levels of seniority. As this was a descriptive exploratory study; a sample size calculation was not performed.

### Study Procedure

Recruited clinicians were block randomised (13) by the primary researcher into one of four groups (Supplementary Table 1) using an online random team generator. Group allocation determined the order in which each participant would complete two sets of five prescribing scenarios (Set 1 and Set 2) and the order of study arms (control or intervention). Prescribing scenarios (Supplementary Table 2) were created by a multi-disciplinary team of clinicians to test a range of common prescribing skills for both adult and paediatric patients. These scenarios included requirements such as ideal body weight calculation, body surface area calculation, route-specific dosing, and a maximum dose limit. Less commonly prescribed medications were used to reduce use of memory and encourage prescribers to use clinical resources and calculate doses where necessary.

Prescribing sessions were recorded using a high-definition camera coupled with desktop screen recording to aid collection of timing and workflow data that would not otherwise be feasible to collect in real-time. All participants viewed a four-minute introductory training video of the intervention, followed by completing two practice scenarios. These were completed with assistance of the researcher if the participant asked for guidance.

Medication scenarios were presented to the participants on paper along with relevant patient and clinical information, e.g., patient sex, age, weight, indication for use, relevant medical history. Participants were asked to prescribe five medications using the intervention and five using the usual resources available to them at the trust. After a dose was determined, participants manually entered the medication order for the test patient on the Cerner Millennium PowerChart (10).

Following completion of each study arm, participants were asked to complete a NASA Task Load Index (TLX) survey (14). Upon completion of both study arms, they were invited to take part in a brief semi-structured interview. This included questions about their experience using the intervention, potential future features and how they perceived it could be integrated into practice (Supplementary Information 1).

### Outcome definitions

Definitions for each outcome measure, adapted from previous work (13), are summarised below.

#### Erroneous medication orders and prescribing errors

An ‘erroneous medication order’ was defined as a medication order associated with one or more prescribing errors. A prescribing error was anything that deviated from recommendations in BNF, BNFC and/or trust guidelines. Prescribing errors could comprise one or more of the following: wrong dosing (deviation >10% outside recommended dosing range), route, frequency, patient, formulation or brand (where relevant). We defined large magnitude dosing errors as deviation >25% outside recommended dosing range.

#### Time to prescribe (TTP)

This was calculated from the time the participant began to read the first scenario, and for subsequent scenarios from the time they completed the previous scenario and moved onto the next. The end point was when the participant submitted the medication order on the eP system.

#### Prescribing workflows

Workflow steps for both the control and intervention arms were created using previously known or anticipated prescribing workflows and adapted if new unanticipated steps were identified during the study observations. These workflows were then utilised for hierarchical task analysis as described in the data analysis section.

### Data collection

Participant demographic information was collected before commencement of the session. The researcher kept field notes to assist with analysis of observations and interviews. NASA-TLX questionnaires were completed by the participant after each of the study arms. Retrospective review of the audio-visual recordings were used to collect the timings, workflow and interview data. All data was entered into an Excel spreadsheet prior to analysis.

Recruitment, consent, randomisation and data collection were conducted by the first author, a female paediatric nurse/researcher and PhD student. She was known to some participants as she was employed in the trust concerned.

### Data analysis

#### Error identification

All potential errors identified by the primary researcher were presented to four pharmacists and medication safety researchers who were blinded to participant and study arm in which the potential error was observed. Each potential error was discussed to determine whether it was in fact erroneous and if so, what error type(s) were present, until consensus achieved.

#### Quantitative analysis

Initial descriptive analyses were conducted on all quantitative outcomes. Univariate and multivariate logistic regression was used to examine the association between the variable of interest (study arm) and the incidence of erroneous medication orders, with and without controlling for the study period and medication set as covariates. Univariate and multivariate quantile regression models were employed to estimate the relationship between the lower, median and upper quartile of the ‘time to prescribe’ and the study arm, again including the study arm, study period and medication set as covariates. Finally, for NASA TLX scores, we calculated mean scores for overall workload and each individual domain per arm, followed by two-sided t-tests to compare mean scores between the control and intervention arms. All statistical analysis was conducted using STATA Version 18 (15).

#### Qualitative analysis

Audio-visual recordings of semi-structured interviews were transcribed verbatim, and reflexive thematic analysis performed on the interview transcripts by the primary researcher, guided by Braun and Clarke’s method (16–18). This method included initial familiarisation of the data by repeated reading of the transcripts, followed by initial code generation. The codes were grouped into themes and the themes were then reviewed and refined, with clear definitions and names assigned.

#### Hierarchical task analysis

Hierarchical Task Analysis (HTA) was conducted by analysing the audio-visual recordings of all erroneous medication orders. The researcher identified where the error appeared to have originated in the prescribing workflow, thus enabling identification of vulnerable workflow steps.

#### Triangulation of outcomes and results

The individual outcomes, methods and analysis were considered collectively, for example erroneous medication orders were mapped onto the HTA workflows and errors are presented in this paper in a way that illustrates not just the incidence of erroneous medication orders but where in the prescribing workflow they occurred/originated. In addition, the results were synthesised collectively to create recommendations for practice and further research, as well as to inform ongoing development and implementation plans for the intervention.

### Registration, approvals and reporting

The study is registered on clinicaltrials.gov (reference NCT05493072) and received HRA approval (IRAS project ID: 315652, REC reference 22/HRA/2896). It is reported using CONSORT and the simulation study extension (19).

## Results

Data were collected during 24 participant sessions, each comprising two sets of 5 simulated medication orders, one based on current practice and one using the intervention. Participants comprised 20 doctors and four pharmacist prescribers (Table 1).

**Table 1.** Participant demographics. PICU-Paediatric Intensive Care Unit, Adult-adult medical or surgical. ST / CT-These abbreviations indicate different stages of speciality training (ST) or core training (CT) in various medical specialities in the United Kingdom. The number following “ST” or “CT” denotes the specific year of training in the speciality or core training program (20).

|  |  |  |  |
| --- | --- | --- | --- |
| **Age** | 25-34 | 10 | 41.7% |
|  | 35-44 | 9 | 37.5% |
|  | 45-54 | 3 | 12.5% |
|  | Not stated | 2 | 8.3% |
| **Gender** | Female | 15 | 62.5% |
|  | Male | 8 | 33.3% |
|  | Not stated | 1 | 4.2% |
| **Profession** | Doctor | 20 | 83.3% |
|  | Pharmacist | 4 | 16.7% |
| **Speciality (self-reported)** | Paediatric Emergency | 1 | 4.2% |
|  | PICU | 4 | 16.7% |
|  | Paediatrics | 9 | 37.5% |
|  | Adult | 10 | 41.7% |
| **Participant Grade (self-reported)** | Foundation Year 1 | 1 | 4.2% |
|  | Senior House Officer | 1 | 4.2% |
|  | ST/CT Years 1-5 | 5 | 20.8% |
|  | ST Years 6-8 | 6 | 25.0% |
|  | Registrar | 1 | 4.2% |
|  | Clinical Fellow | 3 | 12.5% |
|  | Trust grade | 2 | 8.3% |
|  | Consultant | 1 | 4.2% |
|  | Pharmacist Pay Band 8A | 1 | 4.2% |
|  | Pharmacist Pay Band 8B | 3 | 12.5% |
| **Years using Cerner** | < 1 year | 6 | 25.0% |
|  | 1-2 years | 2 | 8.3% |
|  | 2-3 years | 6 | 25.0% |
|  | 3-4 years | 2 | 8.3% |
|  | 4-5 years | 2 | 8.3% |
|  | 5+ years | 6 | 25.0% |

### Prescribing errors

#### Erroneous medication orders

We observed 34 erroneous medication orders in 120 medication orders (28.3%) in the control arm, and eight (6.6%) in 120 medication orders in the intervention arm. Overall error counts by arm, period and medication set are presented in Table 2. The intervention was associated with a 76.5% relative risk reduction (21.7% absolute risk reduction) in erroneous medication orders, demonstrating a statistically significant reduction (odds ratio 0.18; 95% confidence interval (CI) 0.08-0.41; p <0.01). Accounting for both the period and medication set in a multivariate logistic regression model, the intervention is associated with a statistically significantly lower odds of error (OR 0.16, p < 0.01).

**Table 2.** Univariate and multivariate regression outputs for models examining the relationship between use of the intervention (Arm), the covariates (Period and Set) and the incidence of erroneous medication orders. * n = number of prescribing errors and percentage out of the total 120 medication orders observed per arm. **Bold** indicates reference category.

|  |  | Univariate Model |  |  | Multivariate Model |  |  |
| --- | --- | --- | --- | --- | --- | --- | --- |
| Erroneous medication orders | n (%)* | Odds ratio | 95% confidence interval | p value | Odds ratio | 95% confidence interval | p value |
| *Arm* |  |  |  |  |  |  |  |
| **Control** | 34 (28.3%) | 1 | - | - | 1 | - | - |
| Intervention | 8 (6.6%) | 0.18 | 0.08 – 0.41 | <0.01 | 0.16 | 0.06 – 0.43 | <0.01 |
| *Period* |  |  |  |  |  |  |  |
| Period 1 | 12 (10%) | 0.33 | 0.16 – 0.69 | <0.01 | 1.1 | 0.41– 3.18 | 0.79 |
| **Period 2** | 30 (25%) | 1 |  |  | 1 |  |  |
| *Set* |  |  |  |  |  |  |  |
| **Set 1** | 27 (22.5%) | 1 | - | - | 1 | - | - |
| Set 2 | 15 (12.5%) | 0.49 | 0.25 – 0.98 | 0.04 | 0.43 | 0.17 – 1.04 | 0.06 |

#### Prescribing errors by type

In the control arm, route errors were observed in 3.3% of all orders, while no wrong patient errors were observed, as shown in Table 3. Frequency errors were observed in 2.5% of medication orders, and formulation errors in 5.0%. Brand errors were relatively common, accounting for 33.3% of the 12 cases for which the brand was required, and large magnitude errors in 18.3% of cases.

**Table 3.** Number of prescribing errors - by error type. * n = number of prescribing errors and percentage of the total 120 medication orders observed per arm, with the exception of ** brand errors that were relevant to 12 orders per arm where brand specificity was required

| Error | Control<br>(n (%))* | Intervention (n (%))* |  |  |
| --- | --- | --- | --- | --- |
|  |  | Intervention<br>Total | Intervention-<br>external to<br>intervention<br>component of<br>workflow | Intervention-<br>within<br>intervention<br>component of<br>workflow |
| Dose error | 31 (25.8%) | 8 (6.6%) | 7 (5.8%) | 1 (0.8%) |
| Large magnitude<br>dose errors | 22 (18.3%) | 6 (5.0%) | 6 (5.0%) | 0 (0%) |
| Route error | 4 (3.3%) | 3 (2.5%) | 3 (2.5%) | 0 (0%) |
| Patient error | 0 (0%) | 1 (0.8%) | 1 (0.8%) | 0 (0%) |
| Frequency error | 3 (2.5%) | 0 (0%) | 0 (0%) | 0 (0%) |
| Formulation error | 6 (5.0%) | 3 (2.5%) | 3 (2.5%) | 0 (0%) |
| Brand error ** | 4 (33.3%) | 0 (0%) | 0 (0%) | 0 (0%) |

In contrast, in the intervention arm, route errors occurred in 2.5%, and a single patient error was noted (0.8% of orders). Frequency and brand errors were entirely absent, while formulation errors occurred in 2.5%. Large magnitude errors occurred in only 5%.

### Time to prescribe

Two participants, with a total of 20 medication orders, were excluded from TTP analysis due to video recording failure. Therefore, a total of 220 medication orders were included in analysis.

The mean TTP for a medication order in the control arm was 225 seconds (range 57-581), compared to 180 seconds (range 57-480) in the intervention arm. Use of the intervention was not associated with a significant difference in the lower quartile after controlling for the period and medication set (−14 seconds, CI -32.4 to 4.4, p 0.14). There intervention was associated with a statistically significant decrease in the median and upper quartile time to prescribe of 36 seconds (CI -63.9 to -8.1, p <0.01) and 80 seconds (CI -127.2- -32.8 and p <0.01) respectively. Full regression outputs and a box plot are available in Supplementary Tables 3-5 and Supplementary Figure 2.

### Hierarchical task analysis

In the control arm, various steps in the prescribing workflow were identified as causes of error, including failure to identify correct doses, apply maximum dose limits and calculate appropriate dosages based on patient-specific factors. A significant proportion of errors were attributed to Step 3 ‘determine medication and indication’, the step that required prescribers to access and identify the appropriate medication, relevant indication and dose recommendation for the patient (Figure 1 and Supplementary Table 6).

**Figure 1.**
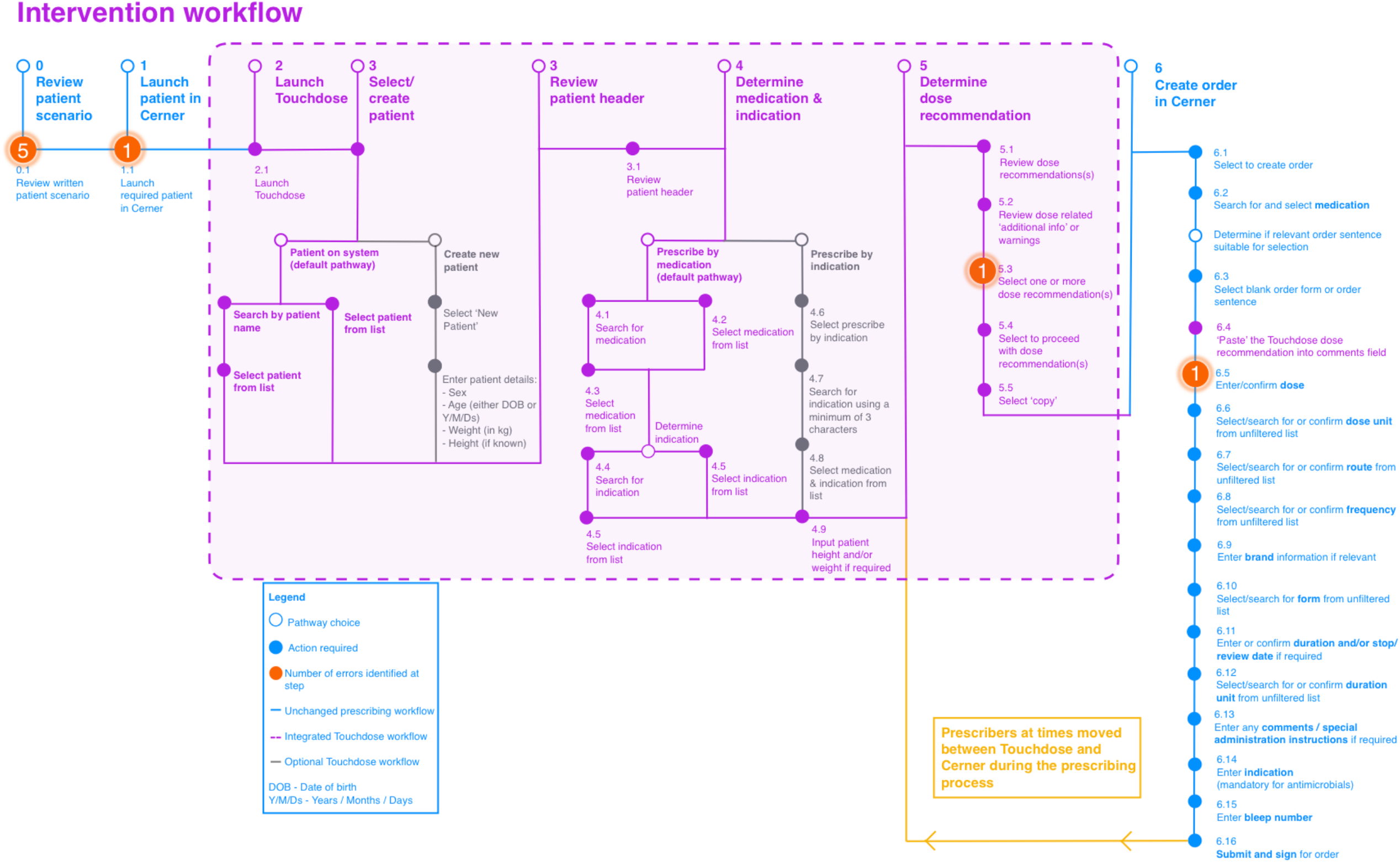
Control arm workflow and hierarchical task analysis identifying erroneous steps observed

In contrast, errors in the intervention arm primarily derived from a failure to identify patient-specific information from the medication scenario (5 of 8 errors). The remaining three errors were a failure to launch the correct patient in Cerner, failure to input a single dose rather than a range, and selection of an incorrect dose for the specified route (Figure 2 and Supplementary Table 7).

**Figure 2.**
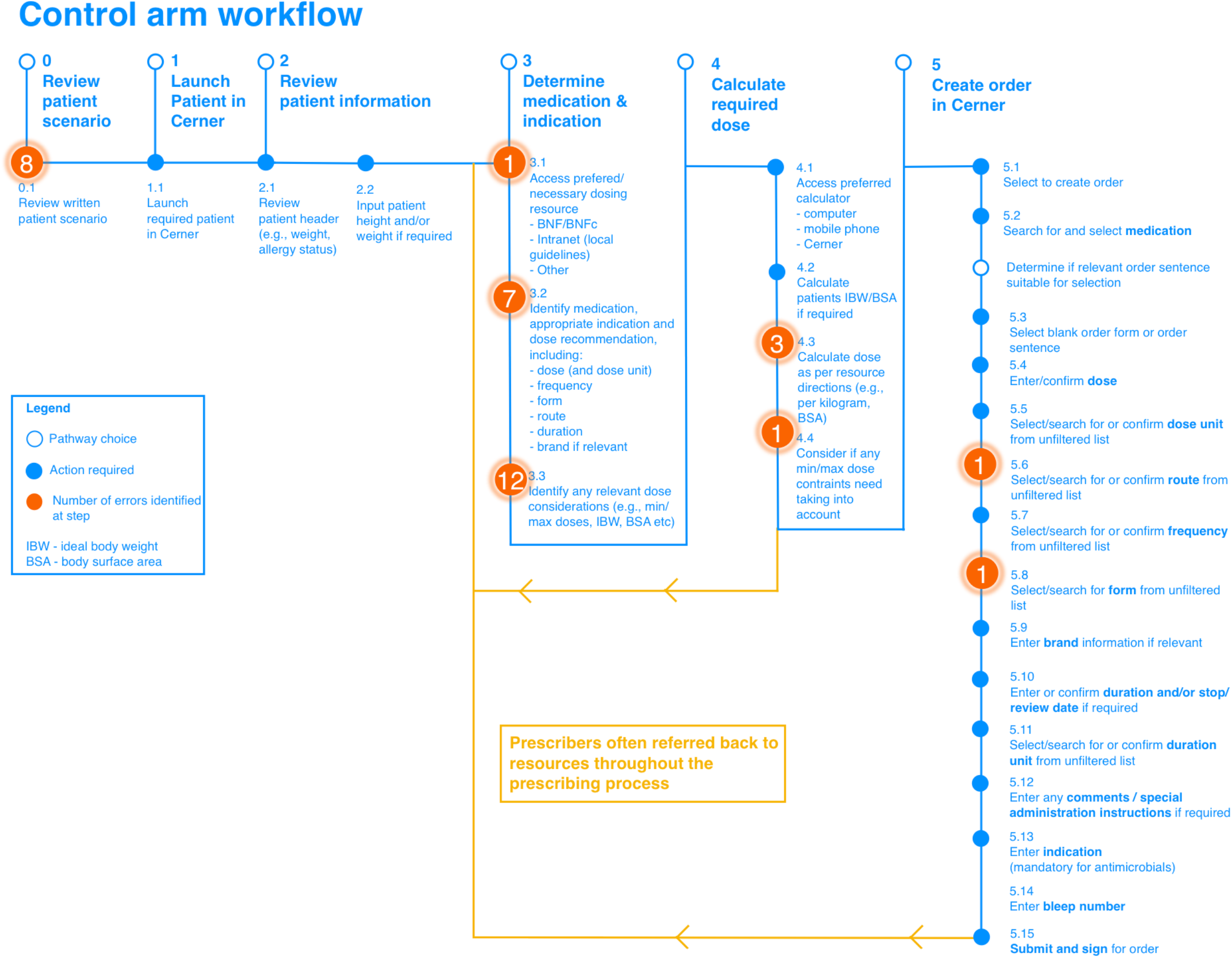
Intervention workflow and hierarchical task analysis identifying erroneous steps observed

### Participant feedback

#### NASA Task Load Index scores

NASA TLX scores revealed that 23 of 24 users perceived a lower task load in the intervention arm compared with control. Two-sample t-tests demonstrated statistically significant differences in overall task load (15.75, p <0.01), mental demand (3.41, p 0.02), temporal demand (2.2, p 0.05), effort (3.88, p <0.01) and frustration (4.54, p <0.01). There was a non-statistically significant trend towards reduction in physical demand and perceived performance. The mean NASA TLX scores and box and whisker plots for each domain by study arm are presented in Supplementary Table 8 and Supplementary Figures 3 and 4.

#### Participant insights

We identified six themes reflecting a variety of participant insights following use of the intervention. These mostly concerned practical considerations that can be used for the continued development of the intervention prior to implementation, prioritisation of future features and proposed patient groups that might most benefit.

*Navigating trust and familiarity: interplay between existing and new systems* Participants expressed mixed experiences with existing systems, citing positive aspects such as locally created care sets and negative experiences due to complexity. Comparisons were drawn between the intervention and other systems; participants noted similarities in functionality but also highlighted that these systems had limitations.

> *“you are not sure [referring to other eP systems] if it’s done it on the right weight or if it’s calculated the body surface or whatever”*
>
> Participant 15
>
> *“What I would say which is really worrying is that these weights are often massively wrong [referring to the weight on the patient header in Cerner]”*
>
> Participant 19
>
> *“have we updated the weight because it could be completely different to what the computer is calculating on and things like that coz that’s what I’ve found sometimes with [system used in intensive care unit at study site]”*
>
> Participant 5

Concerns about trust and reliability of a new system were apparent when articulating known pitfalls of existing prescribing tools/systems. There was therefore a nuanced relationship between trust, familiarity, and reliance on different prescribing resources. Participants expressed a range of sentiments regarding their trust in the intervention versus existing resources such as Cerner and the BNF. Some participants expressed a preference for familiar systems, indicating a need for time to adapt and build trust in new technologies. Others expressed a desire to cross-reference the intervention’s recommendations with the BNF, highlighting a need for reassurance and validation of information. Additionally, questions arose about responsibility, with participants questioning whether errors, were they to be encountered in clinical use, would be attributable to the prescriber or to the intervention.

> *“I think that the biggest thing was trusting that the information was being pulled over correctly”*
>
> Participant 8
>
> *“Sorry I’m just more used to Cerner”*
>
> Participant 12

##### Addressing challenges and suggestions for improvement in intervention integration

Participants highlighted various negative experiences and made new feature suggestions that would improve use of the intervention, particularly in comparison to existing systems like Cerner and the BNF. Participants expressed concerns about the reliability of Cerner’s weight inputs and the potential for errors when transcribing information from the intervention interface to Cerner’s medication order form.

> *“because you have to transfer the information manually I think I feel this is quite like error prone”*
>
> Participant 7

Additionally, there were suggestions for improvements, such as providing renal and hepatic function warnings and adjustments, allergies and pregnancy status. Concerns also arose about technical issues such as wireless internet connectivity, and usability issues such as small text size and unclear navigation.

##### Integration of local guidelines and existing CDSS

Participants enquired about how the intervention will integrate with existing care sets, indicating an assumption of seamless compatibility with established Cerner workflows. Discussions also revolved around the utility of order sentences and power plans, especially for specific drugs such as vitamin D, highlighting potential for the intervention to generate tailored recommendations based on patient-specific inputs. Additionally, participants expressed a preference for links to local guidelines over the BNF for antimicrobial prescribing, emphasising the importance of aligning with institutional practices.

> *“I think I’d still have to check this [local guideline] unless when I typed in conjunctivitis it popped up with like a first-choice therapy box and a secondary therapy box”*.
>
> Participant 1

When participants were informed of the intervention developer’s intention to integrate the local guidelines for certain medications this was received favourably:

> *“that would definitely be super helpful”*
>
> Participant 17
>
> *“especially if you have a link even for like more cautious one [prescriber] I can see the trust guideline it I guess like save me from having to like find the right guideline which itself can be quite tricky”*
>
> Participant 20

*Intervention endorsement: Enhancing Safety, Efficiency, and User Experience* Participants overwhelmingly highlighted a positive experience and provided endorsement of the intervention system regarding various aspects, particularly in terms of safety, efficiency, and ease of use. Participants appreciated the system’s potential to reduce prescribing errors, streamline workflows by providing all necessary information in one place, and remove the need for complex calculations such as for body surface area. They also praised the user-friendly interface and layout, noting clarity, ease of navigation, and better presentation of BNF data compared to current methods.

##### ‘Search by indication’

Participants generally approved the concept of ‘search by indication’, a future feature of the intervention that was introduced during the semi-structured interview. Most recognised its potential utility, especially for exploring alternative treatment options and navigating through complex medication choices. Participants highlighted scenarios where searching by indication first (rather than medication), for medication groups such as antimicrobials or anti-emetics, could further enhance clinical decision-making and improve patient care. However, there were caveats and uncertainties expressed by some participants. Concerns were raised about the need for alignment with local guidelines, the preference for searching by medication rather than indication, and the reliance on other resources such as the app used locally for antimicrobial prescribing guidelines.

##### Targeting specific patient and staff groups

Many patient groups that might most benefit from the intervention were suggested; these included paediatrics, the elderly, and adults post-heart attack or kidney transplant, those with renal or hepatic failure, breastfeeding, or with obesity. The most frequently mentioned was paediatrics (eight participants), mainly due to the high frequency of patient-specific dose calculations. Staff groups that might benefit included “all doctors” with many specifying junior doctors in particular. Nurse prescribers, intensive care doctors and prescribing pharmacists were mentioned as groups for whom the intervention might be less applicable due to specific needs or roles.

> *“staff groups… I mean most would probably benefit it’s a bit more streamlined particularly for things you don’t prescribe frequently”*
>
> Participant 22
>
> *“I think it would be useful everywhere to be honest… like having tools like this are always helpful”*
>
> Participant 15

## Discussion

This study investigated the efficacy of a patient-specific, indication-based prescribing tool in reducing prescribing errors, improving prescribing efficiency, and alleviating user workload compared to standard practice. The results show a substantial reduction in prescribing errors and median and upper time to prescribe (TTP) quartiles when using the intervention. The HTA identified workflow vulnerabilities related to errors. The intervention mitigated many error types seen in the control arm by streamlining access to patient-specific information, automating dose calculations, and providing clear dose recommendations. However, challenges remained in broader prescribing workflows, such as correctly launching the patient in the electronic health record system and transcribing the correct dose for specific routes of administration. User feedback and NASA TLX scores confirmed the intervention’s positive impact on user experience and workload.

This study’s findings align with growing evidence that indication-based prescribing systems can reduce prescribing errors and improve efficiency (7,21–23). Our results closely match those from Garabedian et al. (7) particularly in terms of error rates and task time when using these systems versus standard practices. Despite differences between US and UK healthcare systems, these combined findings support the adoption of indication-based prescribing systems across various healthcare settings.

As for the earlier US study (7), this study provides further evidence that demonstrates the potential of indication-based prescribing tools in a simulation environment. The positive feedback from participants suggests a higher likelihood of acceptance in clinical settings, as predicted by technology acceptance models (24–26). Ongoing user feedback will be essential for refining future prototypes and ensuring successful implementation across diverse settings and patient groups.

A key aspect of our user-testing process was identifying system and workflow vulnerabilities, which should lead to further error reduction. Similar studies on medication-related prescribing and administration guidance have shown the effectiveness of this approach (27,28). However, small or large changes to individual interventions alone may not ensure widespread adoption of these tools. According to Schiff et al. (29), larger scale, “radical change” and clinician buy-in are necessary. We propose that buy-in at all levels—from patients to prescribers, senior management, and procurement teams—is crucial for implementing and scaling these systems. Given the increasing demands on healthcare services (30), engaging with cautious senior management will require robust evidence to support new interventions.

### Strengths and limitations

The study’s strengths lie in its broad medication scenario selection, diverse participant pool, objective outcome measures, comprehensive analysis methods, and robust statistical analysis. The use of a concurrent triangulation design method allowed for the collection of data and analysis using a combination of methods over a shorter period of time compared to a sequential approach (8). This is the first evaluation of its type in England of a patient-specific, indication-based prescribing tool, and aligns with similar work from the US (7,23).

However, the study also faced limitations, including the simulation setting’s inability to fully replicate real-world clinical complexities. Additionally, the relatively small sample size of 24 participants, from one organisation, may limit generalisability.Future research should address these limitations.

### Recommendations for research and practice

Based on the study findings, we make several recommendations in relation to this intervention. There is a need for further refinement of the interface and deeper integration with the electronic health record. This could help mitigate risk of wrong-patient errors. Additionally, incorporating the ability to ‘push’ the final dose recommendation directly into the medication order form could reduce transcription errors. Real-world evaluations will be crucial to assess the intervention’s impact on clinical outcomes and prescriber workflows. These evaluations should involve diverse healthcare settings and patient populations to ensure generalisability and scalability. More broadly, CDSS tools should integrate with local prescribing guidelines to ensure alignment with institutional protocols and to help clinicians make informed decisions based on local best practices. Development of comprehensive training is essential to ensure that clinicians are proficient at using new interventions, training should include system navigation, interpretation of recommendations and how to integrate the system into existing workflows to maximise user adoption and minimise errors.

### Conclusion

This study demonstrates the potential of an indication-based, patient specific prescribing tool, to reduce error, improve efficiency, and reduce user workload in healthcare settings. The findings underscore the importance of integrating human factors and usability engineering principles into the development of CDSS to optimise user experience and effectiveness. Indication-based, patient-specific prescribing tools represent a promising advancement in medication safety technology, with implications for enhancing patient care and healthcare system efficiency.

## Supporting information

CONSORT

## Data Availability

All data produced in the present study are available upon reasonable request to the authors

