## Supplementary for "Comparing safety, performance and user perceptions of a patient-specific indication-based prescribing tool with current practice: A mixed-methods randomised user testing study"

### Online Supplementary Information

Supplementary Figure 1 – Concurrent triangulation methods applied to this study-

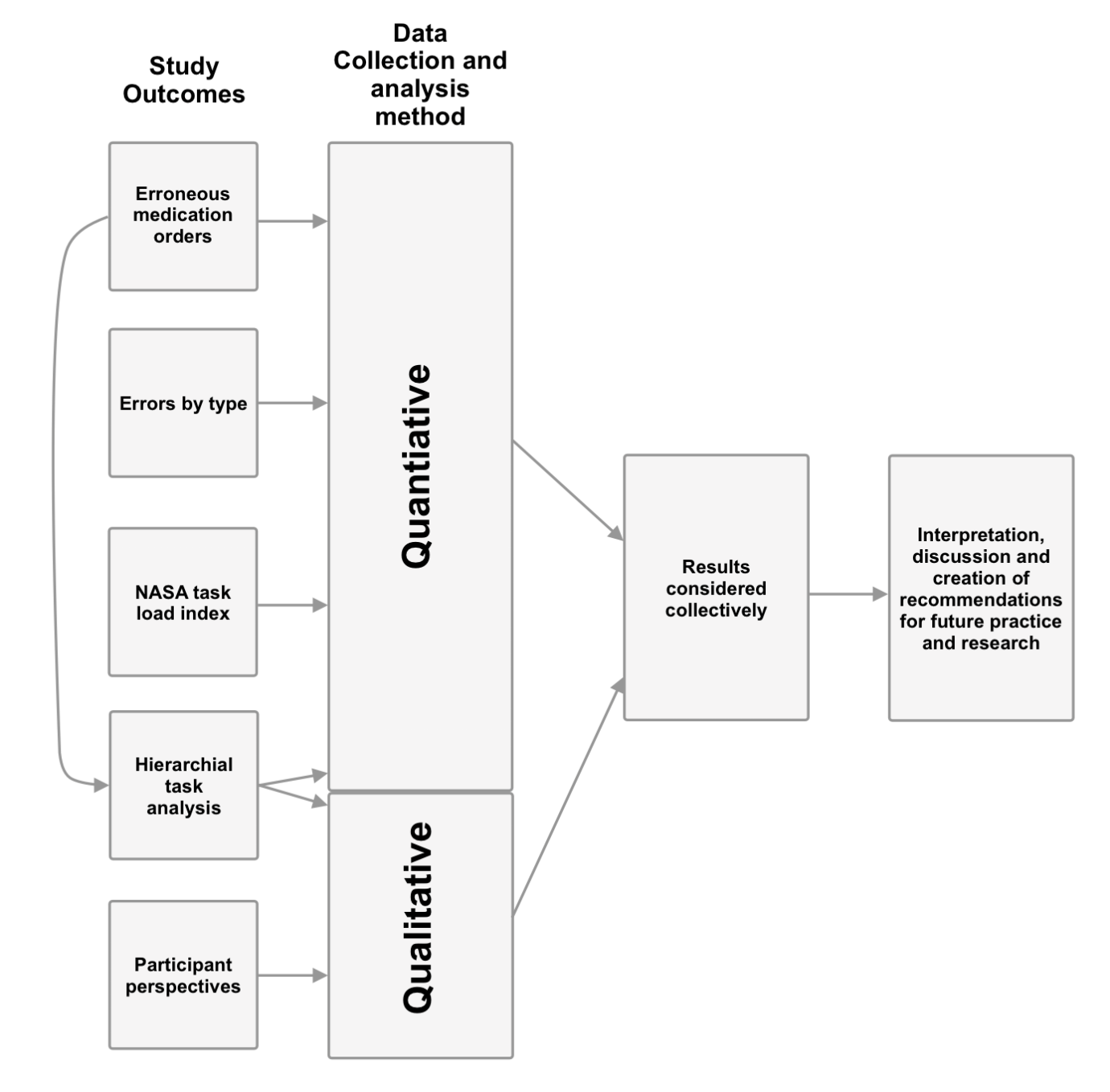

Supplementary Table 1 Block randomisation groups

|  | Group 1 (n = 6) | Group 2 (n = 6) | Group 3 (n = 6) | Group 4 (n = 6) |
| --- | --- | --- | --- | --- |
| Period 1 | Set 1  Control | Set 2  Intervention | Set 1 Intervention | Set 2 Control |
| Period 2 | Set 2  Intervention | Set 1 Control | Set 2  Control | Set 1  Intervention |

n = number of participants randomised to each group that participated in the study

Supplementary Table 2 Simulated scenarios for Group 1 (order of medication set and intervention arm determined by group)
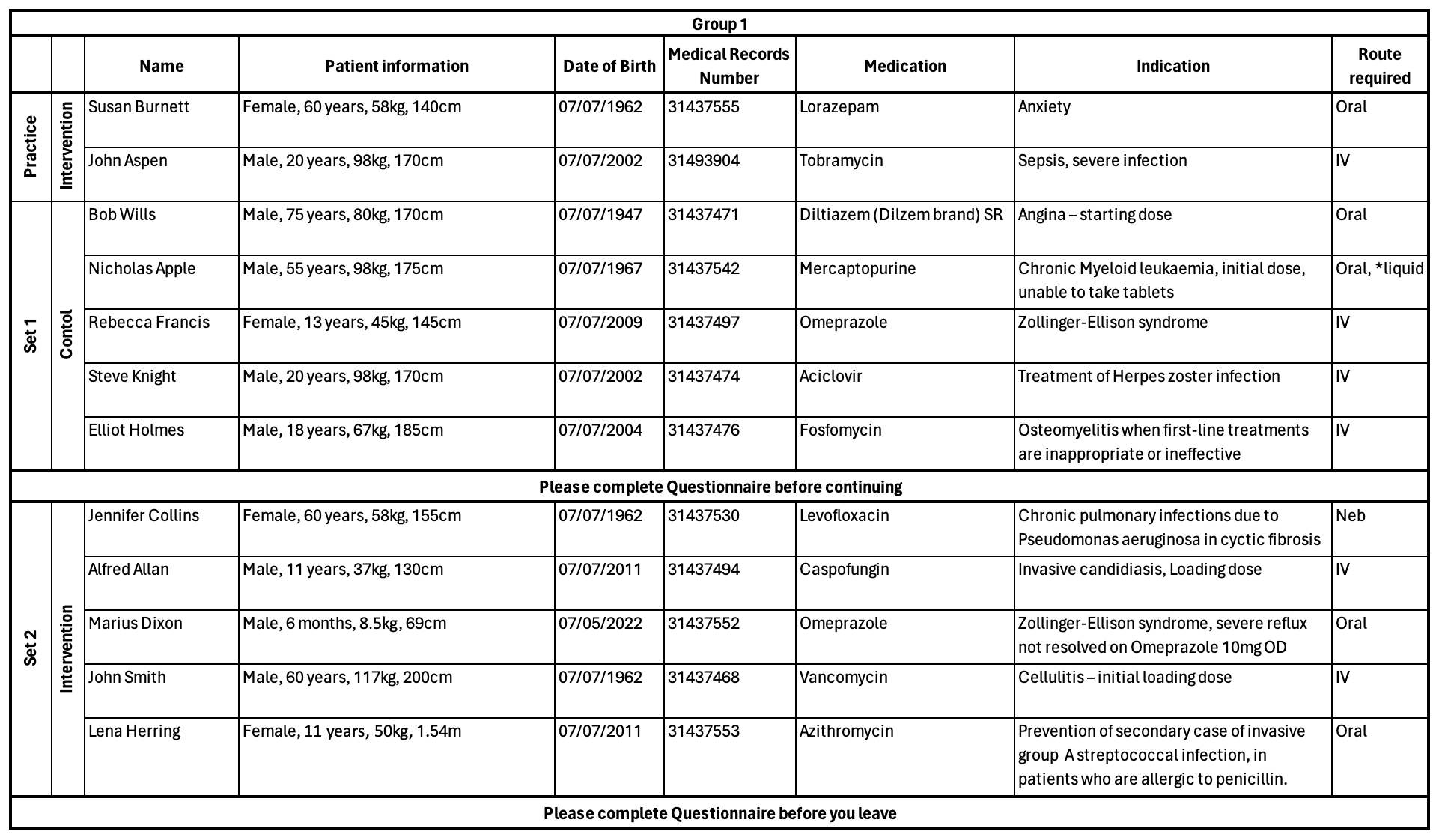
Legend: kg- kilograms IV - intravenous, Neb- nebuliser, OD- once daily

### *Supplementary information 1 - Semi-structured interview topic guide*

Touchdose user testing questions-

- Please tell me how you found today’s user testing session.
  - Could you describe how you found using Touchdose to complete the prescribing tasks.
  - How did this compare with your current prescribing practice?
- What effect do you think a system such as Touchdose could have on your prescribing practice?
  - If positive, in what way?
  - If negative, in what way?
  - Is there anything that you think would benefit from changing/improving?

Indication-based prescribing workflow questions –

- Did you utilise the option to search by indication as opposed to by medication first?
- How does this align with your current prescribing workflow, both your mental workflow and decision making as well as the physical act of prescribing, for example entering the information onto the prescription?
- Would you consider an indication-*first* prescribing workflow to be better/worse/the same as current prescribing workflows. For example, searching for an indication and then selecting a medication, rather than searching for the medication.

Probes –

- Can you expand on that
- Do you have any examples –
  - where this would have been useful...?
  - where this may have hindered...?
  - specific patient groups
  - specific medication types – antimicrobial stewardship

Supplementary Table 3 Univariate and multivariate lower (25^th^) quantile regression outputs for models examining the relationship between the variable of interest (Arm), the covariates (Period and Set) and time to prescribe

|  | Univariate Model | | |  | Multivariate Model | | |
| --- | --- | --- | --- | --- | --- | --- | --- |
| Time to prescribe *25^th^ Quartile regression* | Coef (s) | 95% confidence interval | p value |  | Coef (s) | 95% confidence interval | p  value |
| *Arm* |  |  |  |  |  |  |  |
| **Control** |  |  |  |  |  |  |  |
| Intervention | -17 | -35.07 – 1.07 | 0.07 |  | -14 | -32.40 – 4.40 | 0.14 |
| *Period* |  |  |  |  |  |  |  |
| Period 1 |  |  |  |  |  |  |  |
| **Period 2** | 14 | -3.03 – 31.03 | 0.11 |  | 11 | -7.4 – 29.40 | 0.24 |
| *Set* |  |  |  |  |  |  |  |
| **Set 1** | -17 | -35.59 – 1.58 | 0.07 |  | -11 | -29.33 – 7.33 | 0.24 |
| Set 2 |  |  |  |  |  |  |  |
| Coef – coefficient, (s)- seconds. **Bold** indicates reference category. | | | | | | | |

Supplementary Table 4 Univariate and multivariate median quantile regression outputs for models examining the relationship between the variable of interest (Arm), the covariates (Period and Set) and time to prescribe

|  | Univariate Model | | |  | Multivariate Model | | |
| --- | --- | --- | --- | --- | --- | --- | --- |
| Time to prescribe *Median quartile regression* | Coef (s) | 95% confidence interval | p value |  | Coef (s) | 95% confidence interval | p  value |
| *Arm* |  |  |  |  |  |  |  |
| **Control** |  |  |  |  |  |  |  |
| Intervention | -35 | -62.42 – -7.54 | 0.01 |  | -36 | -63.87 – -8.12 | <0.01 |
| *Period* |  |  |  |  |  |  |  |
| Period 1 |  |  |  |  |  |  |  |
| **Period 2** | 19 | -5.77 – 43.77 | 0.13 |  | 15 | -12.88 – 42.88 | 0.29 |
| *Set* |  |  |  |  |  |  |  |
| **Set 1** | -17 | -43.21 – 9.2 | 0.20 |  | -21 | -48.76 – 6.76 | 0.14 |
| Set 2 |  |  |  |  |  |  |  |
| Coef – coefficient, (s)- seconds. **Bold** indicates reference category. | | | | | | | |

Supplementary Table 5 Univariate and multivariate upper (75^th^) percentile regression outputs for models examining the relationship between the variable of interest (Arm), the covariates (Period and Set) and time to prescribe

|  | Univariate Model | | |  | Multivariate Model | | |
| --- | --- | --- | --- | --- | --- | --- | --- |
| Time to prescribe  75^th^ Quartile regression | Coef | 95% confidence interval | p value |  | Coef | 95% confidence interval | p  value |
| *Arm* |  |  |  |  |  |  |  |
| **Control** |  |  |  |  |  |  |  |
| Intervention | -56 | -103.75 – -8.25 | 0.02 |  | -80 | -127.17 – -32.82 | <0.01 |
| *Period* |  |  |  |  |  |  |  |
| Period 1 | 36 | -7.88 – 79.88 | 0.11 |  | 43 | -4.17 – 90.17 | 0.07 |
| **Period 2** |  |  |  |  |  |  |  |
| *Set* |  |  |  |  |  |  |  |
| **Set 1** | -17 | -63.20 – 29.20 | 0.47 |  | -18 | -64.97 – 28.98 | 0.45 |
| Set 2 |  |  |  |  |  |  |  |
| Coef – coefficient, (s)- seconds. **Bold** indicates reference category. | | | | | | | |

Supplementary Figure 2 Time to prescribe box and whisker plot comparison by arm

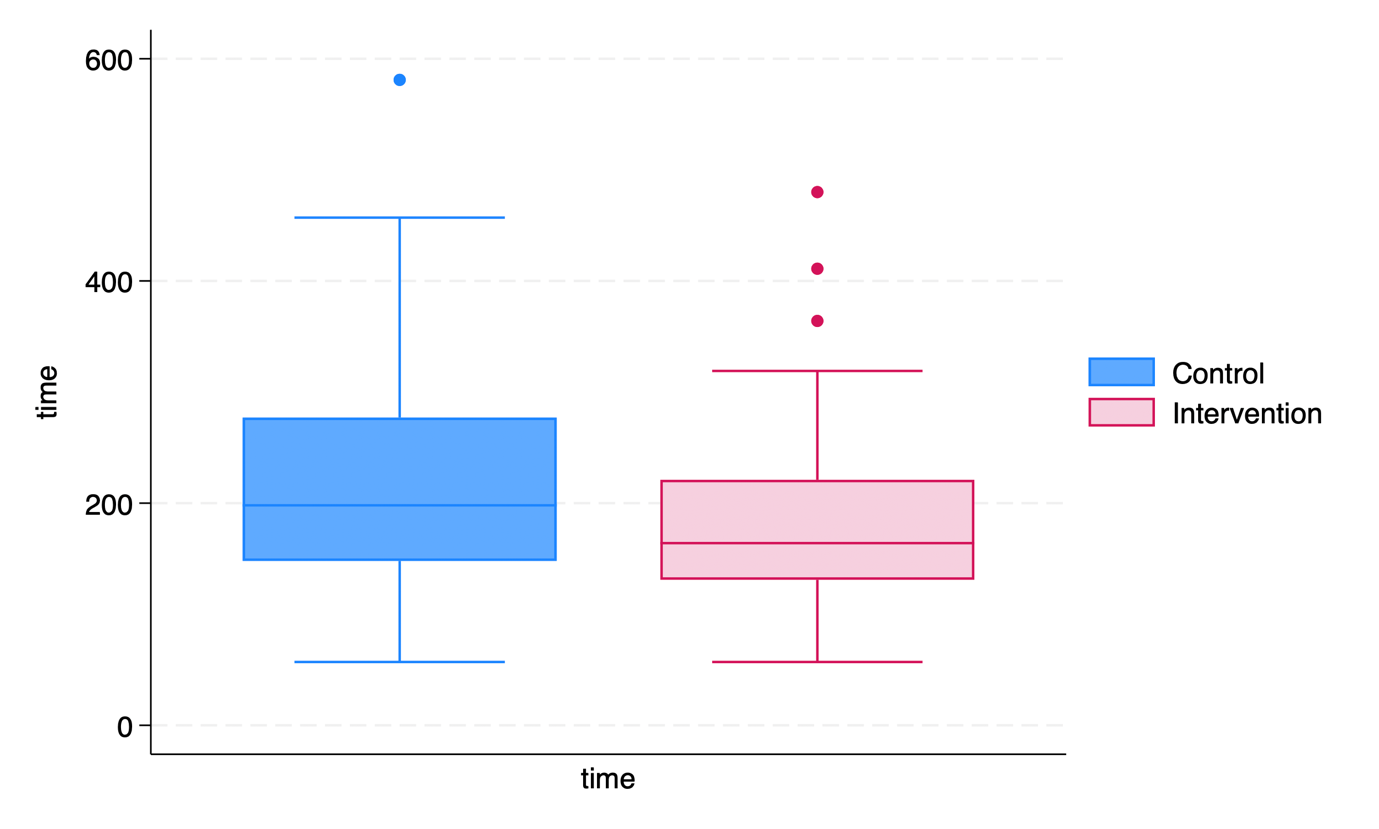

Supplementary Figure 3 Box and whisker plots of NASA Task load index (TLX) overall score across all domains, for control and intervention (Touchdose) arms

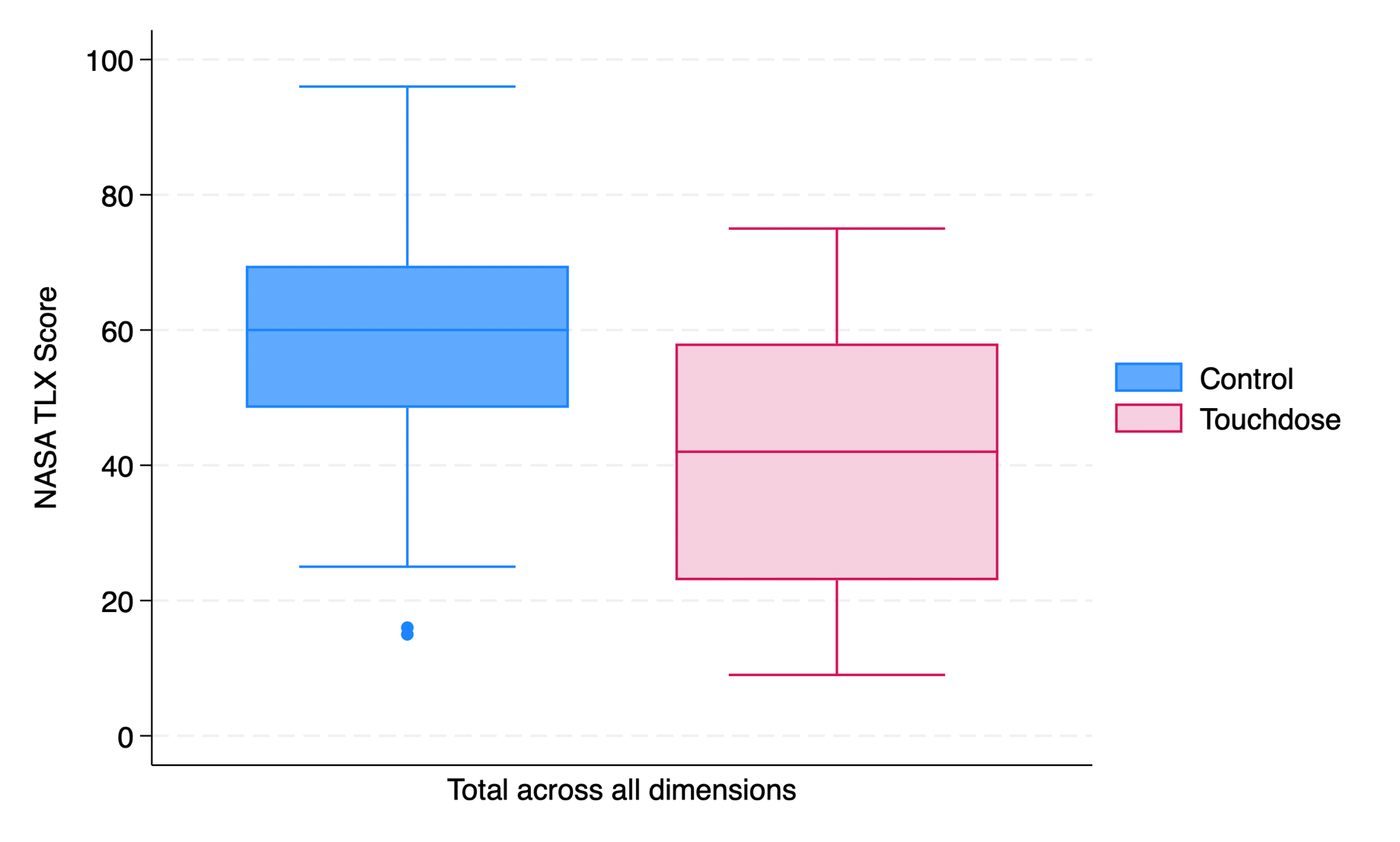

Supplementary 4 Box and whisker plots of NASA Task load index (TLX) scores, by each domain, for control and intervention (Touchdose) arms

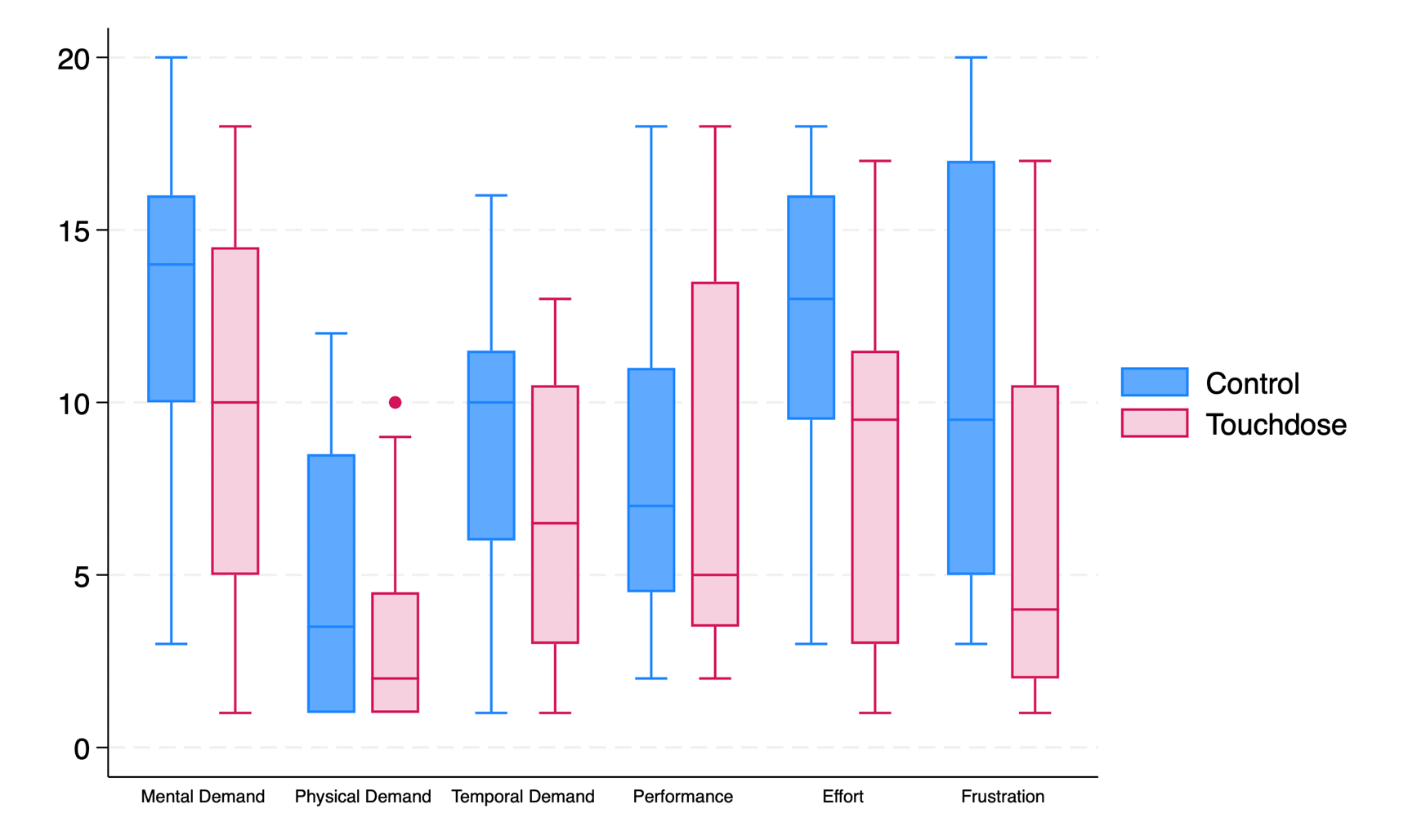

Supplementary Table 6 Hierarchical task analysis for the control arm erroneous medication orders

| Medication | Note | Step(s) associated as contributory factor to error (number relates to workflow flow chart) | Number of occurrences across Control arm |
| --- | --- | --- | --- |
| Aciclovir | Failure to identify dose requires calculation using IBW as patient obese | 3.3/4.4 | 4 |
| Aciclovir | Failure to identify patient specific route requirement on scenario, therefore dose, route, frequency incorrect | 0/3.2 | 1 |
| Aciclovir | Failure to calculate dose based on IBW, inappropriate rounding of dose. (Correctly identified that IBW dose was required) | 4.3/5.3 | 1 |
| Caspofungin | Failure to identify and apply max dose to BSA calculated dose | 3.3/4.4 | 4 |
| Caspofungin | Failure to identify and apply loading dose (lower maintenance dose prescribed) | 0 / 3.2 | 1 |
| Caspofungin | Failure to calculate correct dose using BSA | 4.4 | 1 |
| Caspofungin | Failure to identify and apply max dose to BSA calculated dose | 3.3/4.4 | 1 |
| Diltiazem | Dose for adult patient given rather than elderly patient (patient aged 75years) | 3.2 | 3 |
| Diltiazem | Failure to identify brand specific dosing recommendation, leads to dose, form, brand error | 3.2, 5.4/5.8/5.9 | 1 |
| Diltiazem | Adult dose recommendation transcribed from BNF, using partially completed order sentence for a specific Brand. Therefore dose, brand and frequency incorrect. | 3.2/5.3 | 1 |
| Diltiazem | Failute to identify brand specific dosing recommendation, leads to dose and brand error | 0 / 3.2 | 1 |
| Diltiazem | Failure to identify brand specific dosing recommendation and elderly dose recommendation, therefore dose, frequency and brand error | 3.2/ 5.9 | 1 |
| Fosfomycin | Failure to identify and apply max dose limit | 3.3/4.4 | 1 |
| Levofloxacin | incorrect route, BNF stated inhalation of nebulised solution. order states route inhalation, form solution. | 5.6 | 1 |
| Mercaptopurine | Form error, prescribes tablets instead of oral suspension. Failure to identify patient specific needs from scenario, (it is also the incorrect dose for by tablet) | 0/3.2 | 1 |
| Mercaptopurine | Failure to identify patient specific form requirement on scenario, therefore dose and form incorrect. | 0/3.2 | 1 |
| Mercaptopurine | Selection of a partially completed order sentence with tablet as pre-specified form, failure to edit form to patient specific route requirement. | 5.8 | 1 |
| Mercaptopurine | Failure to calculate dose per meters squared and to input dose correctly ("25mg/m" entered as dose) | 4.3/5.4 | 1 |
| Omeprazole A | Failure to identify patient specific route requirement on scenario, therefore dose, route and form incorrect. | 0/3.2 | 2 |
| Omeprazole A | Using BNF for paediatric patient, prescribes adult dose | 3.1/3.2 | 1 |
| Omeprazole A | Failure to identify and apply route specific dose recommendation | 3.2 | 1 |
| Omeprazole B | Failure to identify and apply max dose limit | 3.3/4.4 | 2 |
| Omeprazole B | Failure to identify patient specific information regarding current dose on scenario, therefore dose error as dose prescribed was less than patients current dose and failure to 'increase if necessary to 20mg' | 0/3.2 | 1 |
| Vancomycin | Failure to calculate correct per kilogram dose | 4.3 | 1 |

Supplementary Table 7 Hierarchical task analysis for the Touchdose (intervention) arm erroneous medication orders

| Medication scenario | Contributory factor to error | Step(s) associated as contributory factor to error (number relates to workflow flow chart) | Number of occurrences across the intervention arm |
| --- | --- | --- | --- |
| Omeprazole A | Failure to identify patient specific route requirement on scenario, therefore dose, route and form incorrect. | 0 | 3 |
| Omeprazole B | Failure to identify patient specific information regarding current dose on scenario, therefore dose error as dose prescribed was less than patients current dose and failure to 'increase if necessary to 20mg' | 0 / (5.3) | 2 |
| Azithromycin | Failure to launch required patient in Cerner (participant prescribed medication for the previous patient). | 1.1 | 1 |
| Mercaptopurine | Failure to input a single dose (participant prescribed dose as a range) | 6.5 | 1 |
| Mercaptopurine | Incorrect dose for route prescribed | 5.3 / (6.5) | 1 |

Supplementary Table 8 NASA-Task load index (TLX) results by arm and TLX domain

| Group |  | Overall task load | Mental demand | Physical demand | Temporal demand | Performance | Effort | Frustration |
| --- | --- | --- | --- | --- | --- | --- | --- | --- |
| Control | Mean | 57.21 | 12.9 | 4.66 | 8.87 | 7.95 | 12.08 | 10.7 |
|  | CI | 48.69 - 65.72 | 10.87 - 14.96 | 3.03 - 6.29 | 7.22 - 10.52 | 6.18 - 9.73 | 10.05 - 14.11 | 8.24 – 13.18 |
| Intervention | Mean | 41.45 | 9.5 | 3.25 | 6.66 | 7.62 | 8.25 | 6.17 |
|  | CI | 33.02 - 49.89 | 7.26 - 11.74 | 2.13 - 4.36 | 5.07 - 8.25 | 5.44 - 9.80 | 6.15 - 10.34 | 3.88 – 8.45 |
| Difference | p | 0.0092 | 0.0244 | 0.1449 | 0.0523 | 0.807 | 0.0092 | 0.0077 |
|  | Mean Absolute reduction | 15.75 | 3.41 | 1.41 | 2.2 | 0.33 | 3.88 | 4.54 |
|  | Relative reduction % | 27.55 | 26.366 | 30.26 | 24.92 | 4.15 | 31.71 | 42.34 |
